# Benchmarking Recording-Level Clinical Speech De-identification for Privacy-Preserving Clinical Audio

**DOI:** 10.64898/2026.04.13.26349911

**Authors:** Hong-Jie Dai, Hang T.B. Le, Liang-Chun Fang, Tatheer Hussain Mir, Ching-Tai Chen, Hui-Hsien Feng, Jiunn-Ru Lai, Hsieh-Chih Hsu, Pratham Nandy, Wei-Hsiang Liao, You-Zhen Tien, Pin-Zhen Chen, Yun-Ru Lin, Jitendra Jonnagaddala

**Author notes:** Corresponding author: Dr. Jitendra Jonnagaddala; Discipline of General Practice, School of Clinical Medicine, UNSW Sydney, Kensington NSW 2052.

## Abstract

Transcript-based de-identification protects textual records but leaves sensitive health information (SHI) directly audible in source recordings, limiting trustworthy reuse of clinical audio. However, standardized evaluations of recording-level SHI detection remain limited. We established the SREDH-AICUP 2025 benchmark to evaluate recording-level SHI detection systems and characterize the approaches. All 22 qualifying systems combined automatic speech recognition with downstream SHI detection, whereas we additionally evaluated end-to-end detection from audio. Pipeline baselines outperformed the end-to-end baseline in transcription (MER, 0.123 vs 0.155) and SHI detection (macro-F1, 0.507-0.653 vs 0.422). Synthetic Mandarin speech-text augmentation enabled the end-to-end model to detect Mandarin SHI that was not identified without augmentation (macro-F1, 0.171 vs 0). Foundation models showed category-specific differences in SHI recognition, and type-aware adaptive routing improved macro-F1 from 0.656 to 0.697. These findings establish standardized reference performance, characterize current system designs, and demonstrate how language-specific augmentation and category-aware model integration can improve recording-level SHI detection.

## Introduction

Spoken clinical interactions are increasingly recognized as a valuable source of healthcare information that complements structured electronic health records (EHRs) and free-text clinical documentation. Doctor-patient consultations, telemedicine encounters, patient interviews, and speech-enabled clinical documentation capture rich contextual information that supports clinical research, automated documentation, clinical decision support, medical education, and healthcare analytics ^1,2^. As advances in artificial intelligence (AI) continue to expand the secondary use of clinical speech, these recordings are increasingly collected, shared, and reused for research and clinical applications ^3,4^. Consequently, protecting sensitive health information (SHI) throughout the clinical speech lifecycle has become essential for enabling trustworthy data reuse and responsible clinical AI.

Protecting SHI has long been recognized as a prerequisite for the secondary use and sharing of clinical data ^5–8^. Over the past two decades, community benchmark initiatives have substantially advanced clinical text de-identification by establishing standardized datasets and evaluation frameworks that enabled reproducible comparison of independently developed methods across diverse clinical documents ^9,10^. These benchmarks not only facilitated compliance with privacy regulations, including the Health Insurance Portability and Accountability Act (HIPAA) ^11,12^ and the European General Data Protection Regulation ^13^, but also accelerated methodological innovation through common evaluation standards. More recently, the rapid emergence of foundation models and generative AI has further increased the demand for reliable privacy-preserving access to clinical data, making robust de-identification an increasingly important component of responsible clinical AI ^14^.

Unlike text-based clinical de-identification, extending privacy protection to spoken clinical interactions fundamentally changes the target of evaluation. Whereas text-based de-identification systems identify SHI as discrete textual entities, recording-level clinical speech de-identification requires systems to detect SHI directly within continuous audio by jointly predicting both semantic categories and temporal boundaries. This recording-level representation provides the temporal information required to selectively remove, redact, or mask sensitive portions of audio while preserving clinically informative content. Existing approaches to audio de-identification have typically combined automatic speech recognition (ASR) with downstream SHI detection and temporal alignment between transcribed text and source recording. Such pipelines, however, can propagate transcription and alignment errors, and the spontaneous nature of clinical conversations can propagate, leading to incomplete localization of SHI spans or unnecessary masking of non-sensitive speech ^15–21^. These characteristics fundamentally distinguish recording-level clinical speech de-identification from conventional text-based de-identification and highlight the need for evaluation methods specifically designed to assess temporally localized SHI detection.

Recent studies have begun to demonstrate the technical feasibility of recording-level speech de-identification by extending sensitive information detection from text to spoken language ^22–25^. In particular, Cohn, et al. ^23^ introduced an audio de-identification benchmark integrating ASR, SHI detection, and audio-text alignment to identify sensitive speech spans within continuous recordings. Their benchmark incorporated annotated segments from the Switchboard ^24^ and Fisher ^25^ conversational speech corpora to support evaluation of audio-level de-identification. However, these resources were derived primarily from general-domain conversational speech and used relatively coarse SHI annotation schemes, limiting their applicability to clinically representative speech scenarios. Meanwhile, existing medical speech corpora have primarily been developed for ASR, clinical dialogue generation, and related conversational AI applications. For example, the corpus introduced by He, et al. ^22^ supports telemedicine dialogue generation but does not include SHI annotations for recording-level de-identification. Consequently, although prior work has established the feasibility of audio-level de-identification, standardized evaluation of clinically oriented recording-level SHI detection remains limited, particularly for comparing independently developed systems, establishing reproducible reference baselines, and evaluating methodological strategies under consistent conditions.

Building on our previously developed SREDH-AI Cup SHI speech corpus ^26,27^, we established the SREDH-AICup 2025 benchmark to enable standardized evaluation of recording-level clinical speech de-identification. Beyond serving as a common evaluation platform, the benchmark integrates independently developed community systems, standardized reference baselines, submitted technical reports, and associated workshop proceedings papers with standardized reference baselines, as well as controlled validation experiments, enabling methodological approaches to be compared under consistent experimental conditions. Importantly, the benchmark incorporates a category-aware, duration-based evaluation framework that jointly assesses SHI classification and temporal localization by quantifying correctly localized, overpredicted, and missed SHI intervals along the audio timeline.

The primary contributions of this study are as follows: (i) Recording-level evaluation for clinical speech de-identification: We establish a category-aware, duration-based evaluation framework that jointly measures SHI classification and temporal localization. By quantifying correctly localized, missed, and overpredicted portions of SHI directly along the audio timeline, this framework captures errors that are not represented by conventional entity-level scoring developed primarily for text. (ii) Characterization of community-developed systems: We review 22 independently developed systems and their standardized technical reports to characterize the ASR strategies, downstream SHI recognition methods, data augmentation, rule-based processing, and temporal alignment approaches used under a common benchmark. We further review nine peer-reviewed studies derived from the benchmark to examine the experimental evidence for these approaches and identify questions not addressed in the published studies. (iii) Standardized architectural comparison and end-to-end augmentation: We establish standardized pipeline and direct end-to-end reference systems to enable comparison of alternative approaches under the same benchmark data and evaluation protocol. We further evaluate synthetic Mandarin speech-text augmentation in the direct end-to-end model, addressing a language condition and architecture not examined in the reviewed community-developed systems and published studies. (iv) Foundation model comparison and adaptive routing: Within the standardized pipeline setting, we compare contemporary foundation models at both the overall and SHI-category levels and evaluate whether category-specific differences in model performance can be exploited through type-aware adaptive routing. Collectively, these contributions provide a standardized basis for evaluating and comparing recording-level clinical speech de-identification systems and for investigating factors that affect SHI detection performance.

## Results

### Benchmark population and evaluation setting

Building on our previously developed SREDH-AICup SHI speech corpus ^28^, we established the SREDH-AICUP 2025 benchmark as a standardized evaluation framework for recording-level clinical speech de-identification through the AI Cup 2025 Medical Speech Sensitive Personal Data Recognition Challenge. The benchmark comprised approximately 20 hours of clinically relevant speech with 7,830 temporally localized SHI annotations across 26 categories. The corpus was partitioned into training (1,539 recordings; 10 h), validation (775 recordings; 5 h), and test (710 recordings; 5 h) sets. Details of corpus development, annotation procedures, and quality assessment have been reported previously ^28^.

Figure 1 summarizes the benchmark datasets and evaluation tasks. The speech data were derived from three sources: the previously developed OpenDeID text de-identification corpus ^29^ adapted for spoken-language use, the dataset for the automated medical transcription corpus ^30^, and medical drama recordings provided by the Taiwan Public Television Service ^31^. The benchmark comprised two tasks. Task 1 evaluated medical speech transcription using a predominantly English corpus supplemented with Mandarin Chinese recordings, whereas Task 2 evaluated recording-level SHI detection using temporally localized annotations across 26 SHI categories. The SHI annotations showed a pronounced long-tailed distribution with substantially more annotations for common categories such as DATE, DOCTOR, and PATIENT than for less frequent categories.

**Figure 1.**
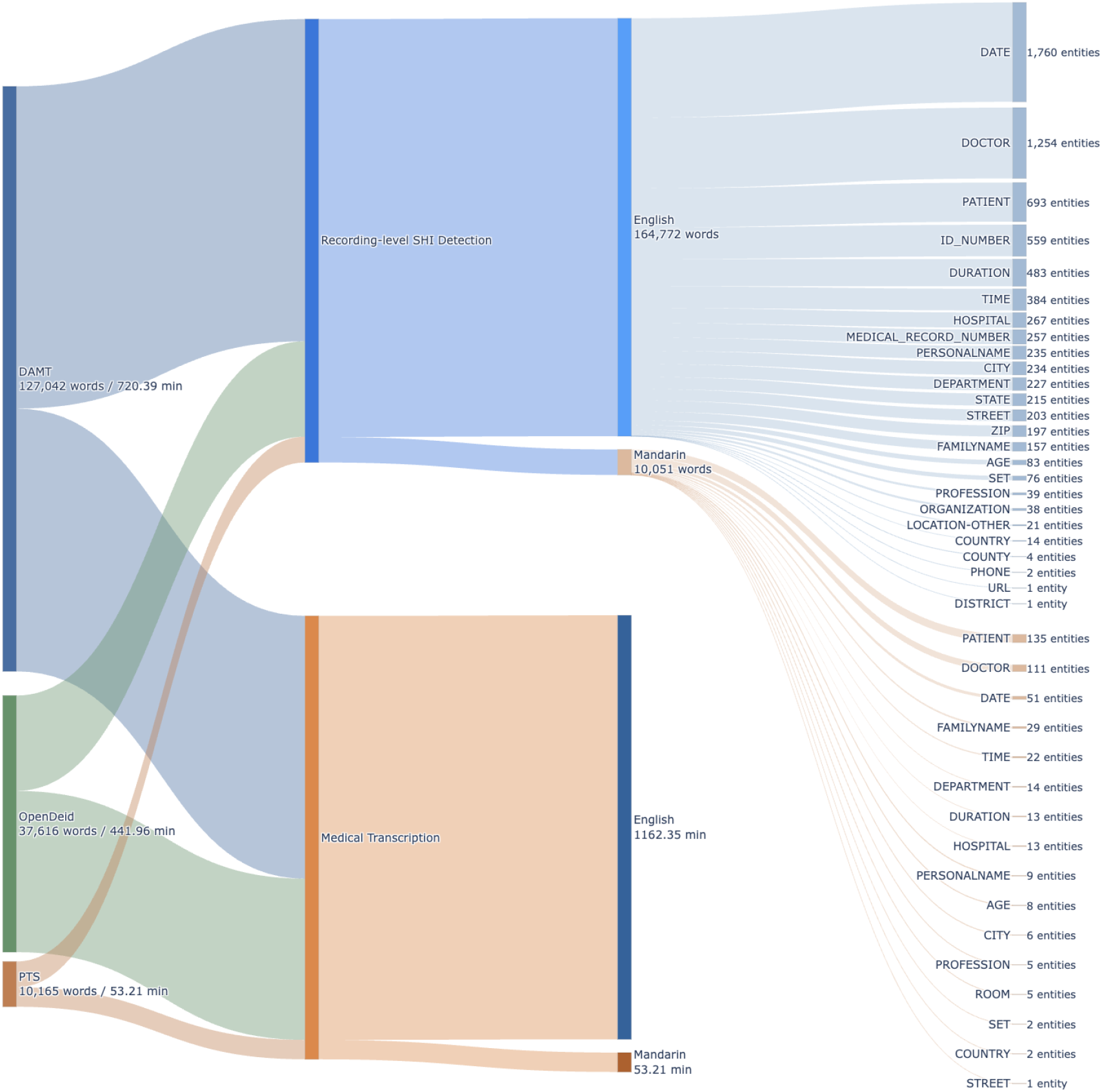
Dataset composition and evaluation tasks of the SREDH-AICUP 2025 benchmark. The benchmark comprises medical speech data derived from the dataset for automated medical transcription (DAMT), OpenDeID, and Public Television Service (PTS) recordings, and includes two tasks: medical speech transcription and recording-level SHI detection. The figure shows the language composition of each task and the distribution of temporally localized SHI annotations across 26 categories.

The benchmark attracted 246 participants forming 149 teams, of which 51 teams submitted valid final predictions, resulting in 160 evaluated submissions. These submissions formed the basis for the subsequent analysis of community-developed systems. All reference experiments reported in this study were evaluated on the same test set using the same task-specific evaluation procedures, enabling direct performance comparison with community-developed systems. The following section presents the standardized reference baselines before examining the methods used by community-developed systems.

### Performance comparison of pipeline and end-to-end baselines

As part of the benchmark, we developed a Whisper-based end-to-end baseline that jointly performed medical speech transcription and recording-level SHI detection. This baseline was also used to define the performance threshold for selecting community-developed systems for subsequent analysis. Because the qualifying community systems used ASR followed by downstream SHI detection rather than direct end-to-end SHI detection, we additionally implemented pipeline baselines for controlled comparison. In these pipeline baselines, transcription and SHI detection were trained separately, and performance was evaluated on the same benchmark test set using the same task-specific evaluation procedures.

We first evaluated pretrained Whisper models ^32^ to determine the transcription configuration for the pipeline baselines (Table 1). Across the pretrained multilingual models, mixed error rate (MER) generally decreased with increasing model size, with relatively small differences among the large-scale Whisper variants. Automatic language detection and manually specified language tokens produced similar MERs across most multilingual models. In contrast, the English-only models achieved lower English MERs for several model sizes but substantially higher Mandarin MERs, resulting in poor overall performance.

**Table 1.**
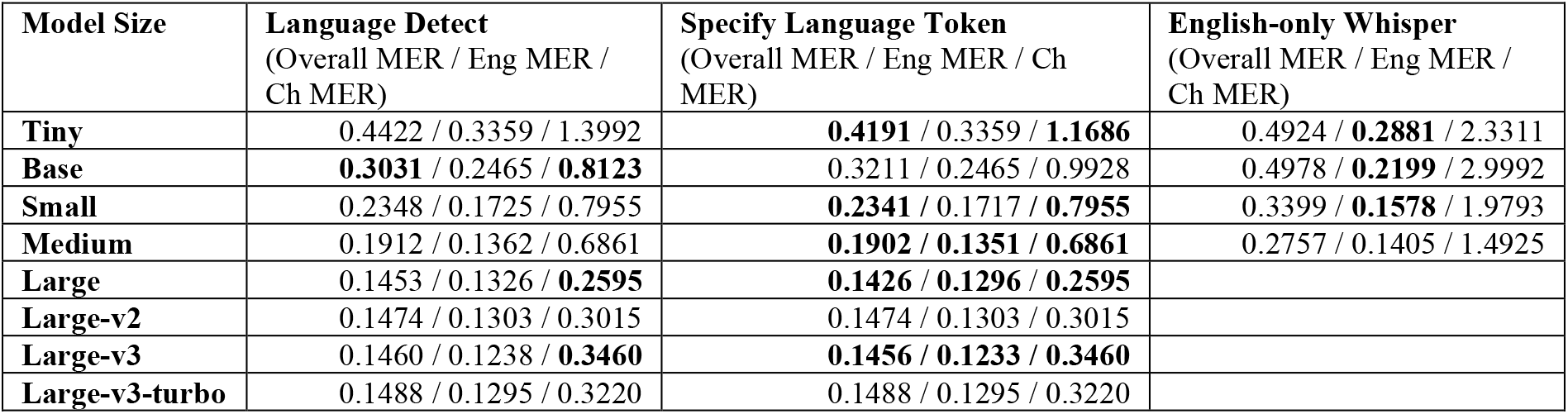
Transcription performance of pretrained Whisper models under multilingual and English-only settings. Mixed error rate (MER) is reported for the overall test set and separately for the English and Mandarin subsets, with lower values indicating better transcription performance. “Language Detect” denotes automatic language identification, “Specify Language Token” denotes transcription using manually specified language tokens, and “English-only” denotes Whisper models trained exclusively on English speech. Boldface indicates the best-performing configuration for each model size.

We next compared bilingual joint training with language-specific independent fine-tuning across Whisper variants (Table 2). The effect of the training strategy varied by model size and language. Language-specific independent fine-tuning substantially reduced overall MER for several larger models, including Whisper large, large-v2, and large-v3, whereas bilingual joint training performed better for some smaller models and large-v3-turbo. Whisper large-v3 consistently outperformed bilingual joint training for larger Whisper models, with the independently fine-tuned Whisper large-v3 achieving the lowest overall MER (0.1227), and was therefore selected as the transcription component of the pipeline baselines for subsequent Task 2.

**Table 2.**
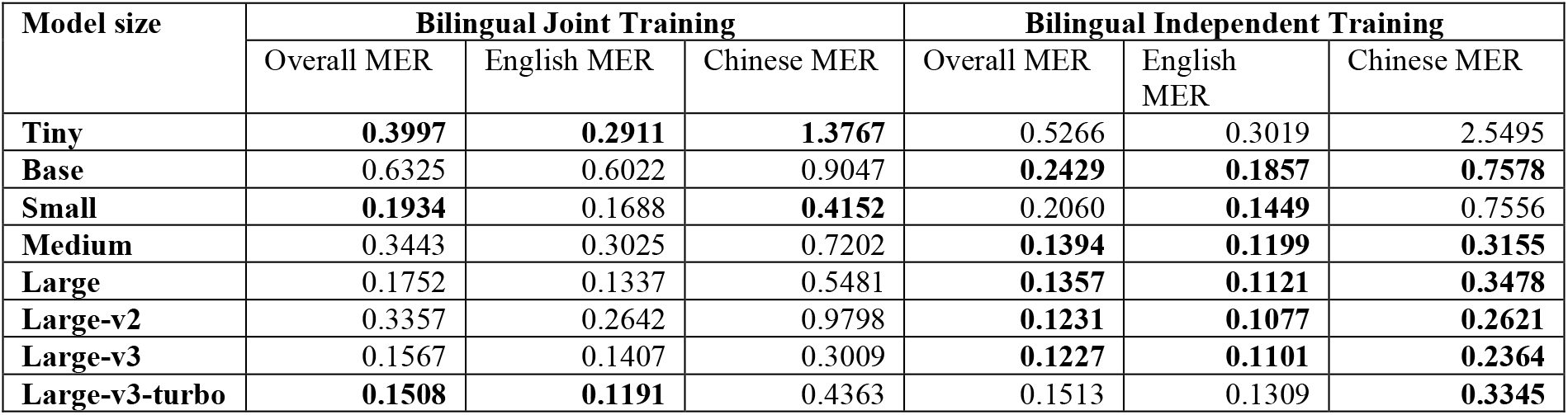
Transcription performance of fine-tuned Whisper models under bilingual joint and language-specific independent training. Mixed error rate (MER) is reported for the overall test set and separately for the English and Mandarin subsets. Bilingual joint training uses a single model trained on both languages, whereas language-specific independent training uses separately fine-tuned English and Mandarin models. Boldface indicates the lower MER between the two training strategies for each model size and evaluation subset.

| Model size | Bilingual Joint Training |  |  | Bilingual Independent Training |  |  |
| --- | --- | --- | --- | --- | --- | --- |
|  | Overall MER | English MER | Chinese MER | Overall MER | English MER | Chinese MER |
| **Tiny** | **0.3997** | **0.2911** | **1.3767** | 0.5266 | 0.3019 | 2.5495 |
| **Base** | 0.6325 | 0.6022 | 0.9047 | **0.2429** | **0.1857** | **0.7578** |
| **Small** | **0.1934** | 0.1688 | **0.4152** | 0.2060 | **0.1449** | 0.7556 |
| **Medium** | 0.3443 | 0.3025 | 0.7202 | **0.1394** | **0.1199** | **0.3155** |
| **Large** | 0.1752 | 0.1337 | 0.5481 | **0.1357** | **0.1121** | **0.3478** |
| **Large-v2** | 0.3357 | 0.2642 | 0.9798 | **0.1231** | **0.1077** | **0.2621** |
| **Large-v3** | 0.1567 | 0.1407 | 0.3009 | **0.1227** | **0.1101** | **0.2364** |
| **Large-v3-turbo** | **0.1508** | **0.1191** | 0.4363 | 0.1513 | 0.1309 | **0.3345** |

Using the Whisper large-v3 transcription configuration for the pipeline, we compared it against end-to-end baselines on the same test set (Table 3). The pipeline baseline achieved a lower overall MER than the end-to-end baseline (0.1227 versus 0.1547). The same pattern was observed for both English and Mandarin, showing that the independently trained transcription component achieved lower transcription error than the joint transcription-SHI model.

**Table 3.** Transcription performance of the official end-to-end ASR-SHI joint training baseline and the independently optimized pipeline baseline. Mixed error rate (MER) is reported for the overall test set and separately for the English and Mandarin subsets. The pipeline baseline uses the language-specific, independently fine-tuned Whisper large-v3 models for transcription, whereas the end-to-end baseline jointly performs speech transcription and SHI detection within a single Whisper-based model. Lower MER indicates better transcription performance.

| Configuration | Overall MER | English MER | Chinese MER |
| --- | --- | --- | --- |
| Pipeline | **0.1227** | **0.1101** | **0.2364** |
| End2End | 0.1547 | 0.1408 | 0.2794 |

We subsequently evaluated recording-level SHI detection using the proposed duration-based evaluation framework (Table 4) in terms of precision (P), recall (R), and F1-score (F). All three pipeline systems achieved higher overall macro-F than the end-to-end baseline (0.507-0.653 vs 0.422) and higher overall micro-F (0.854-0.885 vs 0.812). Among the pipeline models, the Gemma-4 pipeline achieved the strongest overall SHI detection performance across both English and Mandarin subsets. The end-to-end baseline achieved an English macro-F of 0.544 but did not detect SHI in the Mandarin recordings. These results establish the pipeline and end-to-end performance for comparison with the community-developed systems in the subsequent analyses.

**Table 4.** Recording-level SHI detection performance of the pipeline and end-to-end baselines. Recording-level SHI detection performance was evaluated using the duration-based evaluation framework. Macro– and micro-averaged precision (P), recall (R), and F1-score (F) are reported for the overall test set and separately for the English and Mandarin subsets. The pipeline baselines combine the independently fine-tuned Whisper transcription component with XLM-RoBERTa-base, Qwen2.5-Omni-7B, or Gemma-4-E4B-it-qat-q4_0 for downstream SHI detection, whereas the end-to-end baseline jointly performs transcription and SHI detection using Whisper larger-v3. Boldface indicates the highest value within each evaluation metric.

| Method | SHI Detection Model | Overall<br>Macro (P/R/F)<br>Micro (P/R/F) | English<br>Macro (P/R/F)<br>Micro (P/R/F) | Chinese<br>Macro (P/R/F)<br>Micro (P/R/F) |
| --- | --- | --- | --- | --- |
| Pipeline | xlm-roberta-base | 0.546/0.506/0.507<br>0.876/0.836/0.854 | 0.642/0.643/0.634<br>0.882/0.841/0.861 | 0.289/0.315/0.285<br>0.391/0.469/0.426 |
| Pipeline | Qwen2.5-Omni-7B | 0.589/0.562/0.574<br>0.885/0.866/0.875 | 0.757/0.739/0.746<br>0.890/0.876/0.883 | 0.118/0.062/0.079<br>0.258/0.156/0.195 |
| Pipeline | Gemma-4-E4B-it-qat-q4_0 | **0.685/0.656/0.653**<br>**0.893/0.876/0.885** | **0.788/0.761/0.771**<br>**0.902/0.881/0.891** | **0.432/0.510/0.454**<br>**0.400/0.533/0.457** |
| End2End | Whisper-large-v3 | 0.443/0.418/0.422<br>0.848/0.780/0.812 | 0.566/0.556/0.544<br>0.848/0.790/0.818 | 0.000/0.000/0.000<br>0.000/0.000/0.000 |

### Methods used by community-developed systems for recording-level SHI detection

Post-competition surveys completed by 65 participants provided an overview of the methods used by the SREDH-AICUP participants. 62 (95.4%) reported using deep learning-based ASR models, and 53 (81.5%) reported incorporating large language models (LLMs) for downstream SHI detection. Whisper was the most frequently used ASR model family, whereas Qwen ^33^, Llama^34^, and DeepSeek ^35^ were the three most frequently used foundation model families for downstream SHI detection.

Among final submissions, the best Task 1 system achieved an MER of 0.1147, and the best Task 2 system achieved a macro F1-score of 0.7103. Across the best submissions from each team, the mean (median) MER was 0.1687 (0.1650) and the mean (median) macro-F was 0.2760 (0.2980). 19 teams exceeded the end-to-end transcription baseline, and 15 teams achieved a higher macro-F for recording-level SHI detection than the corresponding end-to-end baseline. Based on the predefined inclusion criteria, 22 systems were selected for detailed review.

Table 5 summarizes the main techniques identified from manual review of the 22 systems. All 22 reviewed systems used ASR followed by downstream SHI detection rather than predicting recording-level SHI directly from audio. 17 systems used a single ASR route across recordings, whereas five used language-specific routing with different models or checkpoints for English and Mandarin speech. Whisper and its derivatives were widely used, while the language-specific systems also incorporated alternative ASR models for individual language routes. The resulting transcripts and associated timing information were subsequently used for downstream SHI recognition and for mapping detected SHI spans back to the recording timeline. None of the 22 reviewed systems implemented direct end-to-end recording-level SHI detection without an intermediate transcript.

**Table 5.** Technical approaches identified across the 22 reviewed community-developed systems for recording-level SHI detection. Approaches within each domain were not necessarily mutually exclusive, except for the single-route and language-specific ASR categories. The primary SHI recognition formulation could be determined from the submitted reports for 20 of the 22 reviewed systems; for two systems, the reports identified the LLMs used for SHI recognition but did not provide sufficient information to determine the final prediction formulation. Temporal localization methods could be identified for 19 systems; the remaining three reports did not provide sufficient implementation detail to determine the specific localization method.

| Method Domain | Approach | Systems, n/N (%) | Representative implementations |
| --- | --- | --- | --- |
| ASR | Single-route ASR | 17/22 (77.3%) | One primary ASR route across recordings |
|  | Language-specific ASR routing | 5/22 (22.7%) | Different ASR models or checkpoints for English and Mandarin |
| SHI recognition | Prompt/API-based LLM inference | 7/22 (31.8%) | GPT-3.5/4, Llama, Qwen |
|  | Fine-tuned generative LLM | 6/22 (27.3%) | LoRA/QLoRA adaptation of Gemma, Qwen, Llama, Mistral, or DeepSeek |
|  | Token classification/Sequence labelling | 6/22 (27.3%) | BERT, DeBERTa, and BioLinkBER with BIO/BIOES or CRF |
|  | Custom NER | 1/22 (4.5%) | spaCy NER supplemented with regex |
| Data augmentation | LLM-generated text augmentation | 8/22 (36.4%) | Generation of additional examples for less frequent SHI categories, semantic rewriting, and contextual augmentation |
|  | Entity replacement | 1/22 (4.5%) | Random replacement of selected SHI entities |
| Model specialization and inference | Category-specific models or adapters | 2/22 (9.1%) | Category-specific Qwen models and entity-group-specific QLoRA adapters |
|  | Retrieval-augmented prompting | 2/22 (9.1%) | Dynamic retrieval of labeled examples for in-context prompting |
| Post-processing | Rule- or regex-based components | 10/22 (45.5%) | Entity recovery, lexicon matching, category correction, prediction filtering, and output validation |
| Temporal localization | Explicit localization method | 19/22 (86.4%) | ASR-derived timestamps, WhisperX/forced alignment, and explicit transcript-to-time mapping |
|  | Fuzzy or approximate span matching | 9/22 (40.9%) | Normalized, fuzzy, or Levenshtein-based matching to timestamped words |

For downstream SHI recognition, seven systems used prompt-or API-based LLM inference without final task-specific parameter tuning, six fine-tuned generative LLMs, and six used token classification or sequence labeling approaches, including BIO/BIOES tagging and conditional random fields decoding^36^. One system trained a custom spaCy NER model supplemented with regex-based recognition. For two additional systems, the submitted reports identified the LLMs used for SHI recognition but did not provide sufficient information to determine the final prediction formulation. Beyond the primary SHI detection models, teams incorporated data augmentation, category-specific models or adapters, retrieval of labeled examples, and deterministic processing at different stages of their systems.

Data augmentation was implemented in nine of the 22 reviewed systems. Eight systems generated additional training text using LLMs, whereas one augmented training examples by replacing selected SHI entities. These approaches were primarily used to increase the number or diversity of training examples, including examples for less frequent SHI categories. Reported improvements were not consistent across implementations; for example, one team ^37^ reported that Qwen3-based augmentation reduced F1 by 0.025.

Several systems also adapted downstream processing to differences among SHI categories. For example, one team used seven category-specific Qwen models, whereas another trained six entity-group-specific QLoRA adapters. Two systems used retrieval-augmented prompting to dynamically select labeled examples from the provided training data as in-context demonstrations. Rule-or regex-based components were incorporated by 10 of the 22 systems for entity recovery, lexicon matching, category correction, prediction filtering, or output validation.

Temporal localization was required to convert transcript-level SHI predictions into recording-level outputs. It could be identified from the submitted reports for 19 of the 22 systems. These systems used ASR-derived timestamps, post hoc alignment methods such as WhisperX ^38^, forced-alignment models, or explicit mappings between transcript positions and timestamped words. Nine systems further used fuzzy or approximate span matching, including normalized string matching or Levenshtein-based matching, to associate predicted SHI spans with timestamped transcript segments when exact correspondence was unavailable. For the remaining three systems, the submitted reports did not provide sufficient detail to determine the specific localization method.

Figure 2 compares the standardized benchmark baselines with the union of the 10 highest-performing teams in Task 1 and the 10 highest-performing teams in Task 2. The strongest comparative pipeline baseline achieved an MER of 0.123 and a macro-F of 0.653, whereas the end-to-end baseline achieved an MER of 0.155 and a macro-F of 0.422. Because the official competition ranking was determined by the combined Task 1 and Task 2 ranks, Figure 2 shows the distribution of task-specific performance rather than reproducing the official leaderboard.

**Figure 2.**
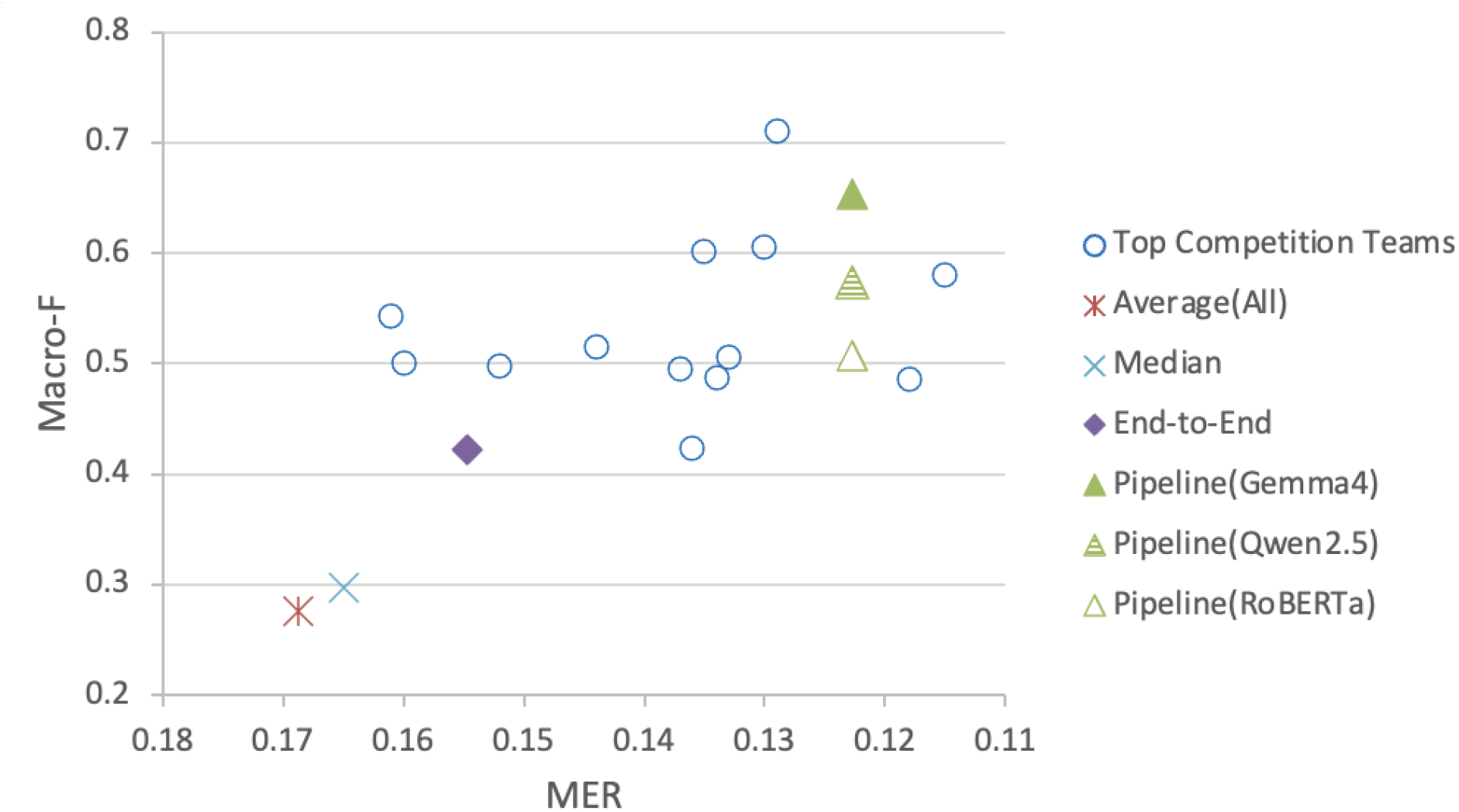
Task 1 and Task 2 performance of representative community-developed systems and standardized benchmark baselines. The scatter plot shows Task 1 transcription performance using MER (lower values indicate better performance) and Task 2 recording-level SHI detection performance using macro F1-score (higher values indicate better performance). Community systems comprise the union of the 10 highest-performing teams in Task 1 and the 10 highest-performing teams in Task 2. Because these groups did not completely overlap, more than 10 unique systems are shown. The end-to-end baseline and comparative pipeline baselines using Gemma4, Qwen2.5, and RoBERTa are shown together with the mean and median performance across final team submissions. The figure shows task-specific performance and does not reflect the official competition ranking, which was determined by the combined ranks of Task 1 and Task 2.

### Experimental evidence from published studies and additional benchmark analyses

Nine peer-reviewed studies derived from the SREDH-AICUP workshop ^39^ were further reviewed to determine which methods identified in the technical reports had been experimentally evaluated and which questions remained unresolved. Across the reviewed studies, six implemented data augmentation, including LLM-generated training examples, acoustic augmentation during ASR training, and lexical resource expansion. All nine reported differences in performance or data availability across SHI categories, and two trained separate models or adapters for specific SHI categories or category groups. However, none evaluated synthetic Mandarin speech augmentation in a direct end-to-end recording-level SHI model, and none examined whether category-specific differences across foundation models could be used for adaptive routing. We therefore conducted two additional benchmark analyses to address these questions.

Data augmentation was used by nine of the 22 reviewed community-developed systems, including systems that specifically augmented Mandarin training data to address its limited representation in the benchmark. We therefore examined the nine published studies to determine how augmentation had been experimentally evaluated. Six of the nine studies implemented augmentation ^37,40–44^, but the approaches differed substantially: three generated additional text examples using LLMs, two applied acoustic augmentation during ASR training, and one expanded lexical or dictionary resources for underrepresented SHI categories. The reported effects were not uniformly positive. One study directly compared models trained with and without LLM-generated text augmentation and reported a decrease in F1-score from 0.5177 to 0.4733 after augmentation; the authors attributed this reduction to mixed-language generation, limited syntactic diversity, and incomplete SHI annotations in the generated examples. Thus, although augmentation was used in both the community-developed systems and published studies, the available evidence did not establish a consistent benefit, and none of the published studies evaluated additional Mandarin speech-text pairs within a direct end-to-end recording-level SHI model.

To address this question, we constructed a Mandarin speech augmentation pipeline, as described in the Methods, to generate additional Mandarin speech-text pairs for end-to-end Whisper training. Following the training strategies established in Table 2, we evaluated the augmented Mandarin speech under both bilingual joint and language-specific independent training. As shown in Table 6, the original end-to-end Whisper-large-v3 baseline detected no SHI temporal spans in Mandarin recordings (Chinese macro F1 = 0.0000). With Mandarin speech augmentation, bilingual joint training increased the Chinese macro F1-score to 0.1706, and language-specific independent training achieved a macro F1 of 0.1411. The two training strategies showed different performance across the English and Mandarin subsets. Bilingual joint training produced higher Mandarin macro-F, whereas language-specific independent training achieved the highest overall and English performance, increasing the overall macro F1-score from 0.422 to 0.439 and the English macro F1-score from 0.544 to 0.566.

**Table 6.**
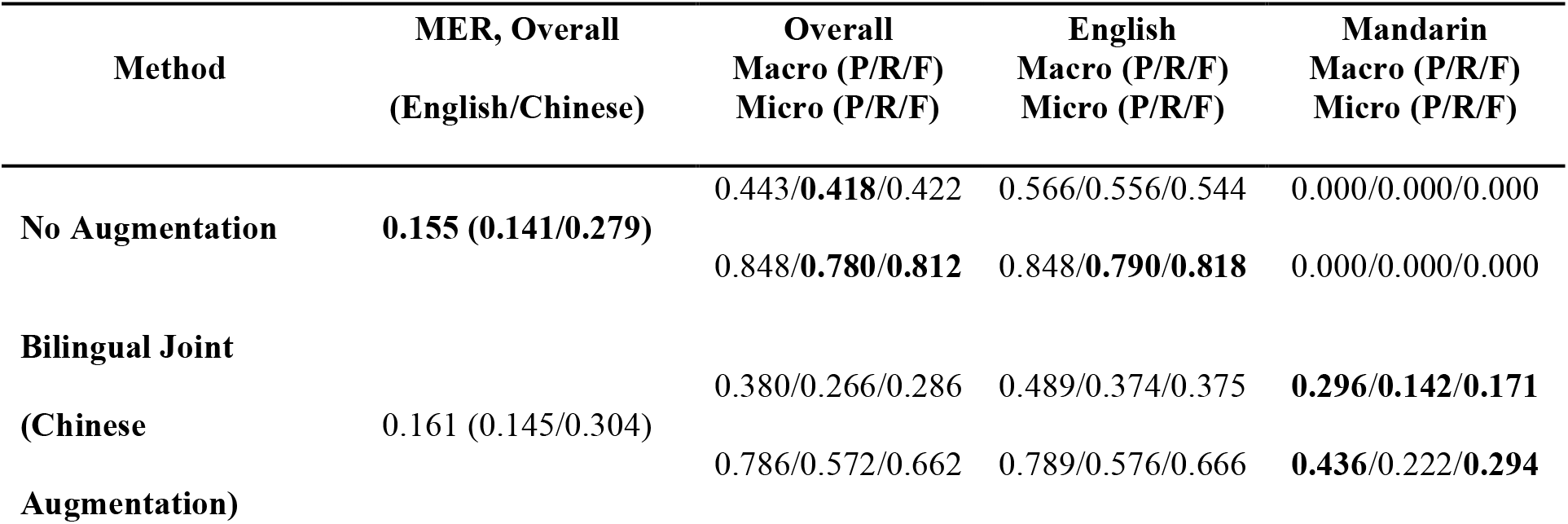
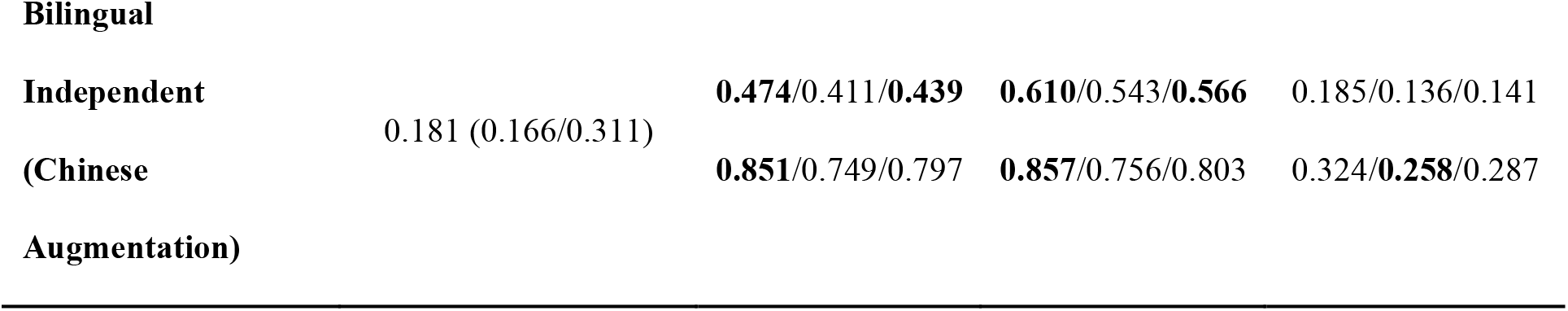
Effect of Mandarin speech augmentation on transcription and recording-level SHI detection under different end-to-end training strategies. Mixed error rate (MER) is reported for the overall test set and separately for the English and Mandarin subsets. Recording-level SHI detection performance is reported as macro– and micro-averaged precision (P), recall (R), and F1-score (F). The no-augmentation condition corresponds to the original end-to-end Whisper-large-v3 baseline. Bilingual joint training uses a single model trained jointly on English and augmented Mandarin data, whereas language-specific independent training uses separately trained models for the English and Mandarin subsets.

The improvement in Mandarin SHI detection was not accompanied by improved transcription accuracy. After removing all predicted <|NER|> tags from the generated outputs, overall MER increased from 0.155 without augmentation to 0.161 with bilingual joint training and 0.181 with language-specific independent training. Mandarin MER likewise increased from 0.279 to 0.304 and 0.311, respectively. Thus, the improvement in Mandarin SHI detection cannot be explained by improved transcription accuracy alone.

Differences among SHI categories were also evident across the nine published studies. All nine reported category-level differences in data availability, recognition performance, or downstream processing, and two explicitly trained separate models ^40^ or adapters ^42^ for specific SHI categories or category groups. Previous work has also highlighted foundation model selection as an important determinant of downstream performance ^45^. However, the reviewed studies did not systematically compare multiple foundation models under the same evaluation setting to determine whether their relative performance differed across SHI categories. We therefore compared six contemporary open foundation models at both the overall and category levels and subsequently examined whether these differences could be exploited through adaptive routing.

We compared four general-purpose models (Qwen2.5-7B, Gemma-2-9B, Mistral-7B, and LLaMA-3.1-8B) and two biomedical models (OpenBioLLM-8B and Med42-8B). For historical comparison, we additionally evaluated the two best-performing baselines based on the Pythia-2.8B used in our previous transcript-level clinical SHI de-identification study ^29^. As shown in Table 7, the six contemporary models substantially outperformed the historical Pythia baseline, increasing micro-F1 from 0.27-0.53 to 0.80-0.87 and macro-F1 from 0.13-0.31 to 0.52-0.66, demonstrating the rapid advancement of recent instruction-tuned foundation models for clinical SHI extraction. Among the contemporary models, LLaMA-3.1-8B achieved the highest micro-F (0.867), whereas OpenBioLLM-8B and Med42-8B achieved higher macro-F scores (0.651 and 0.656, respectively),

**Table 7.** Overall performance and entity-specific strengths of foundation models for clinical SHI recognition. P and R denote overall precision and recall, respectively. Highest-performing SHI categories indicate categories for which each model achieved the highest category-level F1-score among the six contemporary foundation models. Ties are retained; PHONE achieved the same highest F1-score (0.9810) with Qwen2.5-7B, LLaMA-3.1-8B, OpenBioLLM-8B, and Med42-8B. The historical Pythia models were included for overall performance comparison and were not considered when identifying the highest-performing categories. Detailed category-level results are provided in Supplementary Table 1.

|  | Model | P | R | Micro F1 | Macro F1 | Highest-performing SHI categories |
| --- | --- | --- | --- | --- | --- | --- |
| Gen. | Qwen2.5-7B | 0.887 | 0.837 | 0.861 | 0.631 | PATIENT, ZIP, STATE, PHONE |
|  | Gemma-2-9B | 0.857 | 0.746 | 0.798 | 0.517 | PROFESSION, ROOM |
|  | Mistral-7B | 0.885 | 0.839 | 0.862 | 0.599 | DOCTOR, MRN, CITY, STREET, AGE, FAMILYNAME, SET |
|  | LLaMA-3.1-8B | 0.887 | **0.847** | **0.867** | 0.640 | DEPARTMENT, TIME, LOCATION-OTHER, PHONE |
| Bio. | OpenBioLLM-8B | 0.886 | 0.830 | 0.857 | 0.651 | PERSONAL_NAME, ORGANIZATION, ID_NUMBER, PHONE |
|  | Med42-8B | **0.889** | 0.833 | 0.860 | **0.656** | HOSPITAL, DURATION, DATE, COUNTRY |
| His. | Pythia 2.8B | 0.546 | 0.508 | 0.526 | 0.311 | - |
|  | Pythia 2.8B-ICL | 0.384 | 0.212 | 0.274 | 0.130 | - |

No single model achieved the highest F1-score across all SHI categories (Supplementary Table 1), and the highest-performing model varied across categories. We therefore evaluated whether these category-specific differences could be exploited with the type-aware adaptive routing framework described in the Methods section. For each SHI category, candidate models were selected based on their category-specific validation performance, and their predictions were combined using performance-based weights adjusted according to the number of training examples available for that category. The resulting routed model achieved a macro-F of 0.697 compared with 0.656 for the best individual model, with a micro-F1 of 0.863.

## Discussion

This study established a standardized benchmark for recording-level clinical speech de-identification that integrates reference baselines, community-developed systems, and a category-aware, duration-based evaluation framework for temporally localized SHI. Analysis of 22 community-developed systems showed that all used an ASR-to-SHI pipeline, although their downstream SHI recognition, data augmentation, rule-based processing, and temporal alignment methods differed. Under standardized experimental conditions, downstream model selection substantially affected SHI detection performance, highlighting the contribution of this choice to the performance attainable with a common pipeline. Additional experiments evaluated a direct end-to-end formulation not represented among the reviewed community systems and showed that Mandarin speech augmentation improved SHI detection despite increased transcription error, whereas category-level comparison of contemporary foundation models revealed differences that could be further exploited through type-aware adaptive routing. Together, these analyses characterize the methods currently used for recording-level SHI detection, evaluate specific factors associated with performance under standardized conditions, and provide an evaluation framework that accounts explicitly for both SHI category and temporal localization.

The consistent use of ASR-to-SHI pipelines across the 22 reviewed systems indicates that transcript-based processing remained the predominant approach within this benchmark. Whisper and its derivatives were widely used for transcription, whereas downstream SHI recognition included prompt– or API-based LLM inference, fine-tuned generative models, token classification or sequence labeling, and combinations with rule– or regex-based processing. This modular architecture was well-suited to the competition design, in which transcription and recording-level SHI detection were evaluated as separate tasks and could therefore be optimized independently. This may partly explain the consistent adoption of pipeline systems, although the reviewed reports do not establish that pipeline architectures are intrinsically superior to direct end-to-end approaches. Importantly, the objective of recording-level de-identification is not necessarily to produce the most accurate transcript, but to identify sensitive information and determine its location in the source audio. Our direct end-to-end implementation, therefore, provides a complementary formulation that was absent from the reviewed community systems and enables evaluation of recording-level SHI detection without requiring a separately optimized downstream SHI recognition pipeline.

The standardized pipeline experiments further demonstrate the importance of downstream model selection. With the transcription component, dataset split, training and inference settings, and evaluation protocol held constant, substantial differences in SHI detection performance were observed across RoBERTa, Qwen2.5, and Gemma, with the Gemma-based pipeline achieving the strongest performance. Notably, our standardized Gemma-based pipeline did not incorporate several competition-specific refinements observed in the reviewed systems, such as extensive rule-based post-processing or specialized data curation, yet its performance approached that of the highest-ranking community-developed systems. These findings indicate that advances in downstream models can materially raise the performance attainable from a comparatively simple standardized pipeline. This interpretation is also consistent with the substantially higher performance of contemporary foundation models compared with the historical Pythia-2.8B configurations evaluated in our previous transcript-level SHI de-identification framework. Together, these comparisons illustrate that advances in foundation models can increase the performance attainable in clinical SHI recognition, emphasizing the need to interpret benchmark reference performance in the context of the models available at the time of evaluation.

Data augmentation was frequently explored in the published studies, but its effectiveness varied across implementations. Six of the nine reviewed studies used augmentation, including LLM-generated training examples, acoustic augmentation during ASR training, and expansion of lexical resources, and the available evidence did not show a consistent performance benefit. Our experiments extend these previous approaches by evaluating synthetic Mandarin speech-text augmentation in a direct end-to-end model, a setting not examined in the reviewed studies. Augmentation enabled the end-to-end model to detect Mandarin SHI that was not identified without augmentation, although the extent of improvement differed between bilingual joint and language-specific independent training. These findings suggest that the value of augmentation depends on both the data condition and how augmented data are incorporated into model training, rather than augmentation alone being uniformly beneficial.

The improvement in Mandarin SHI detection was not accompanied by an improvement in transcription accuracy. MER increased under both augmentation strategies relative to the original end-to-end model, despite improved SHI detection in the Mandarin subset. This finding does not imply that transcription quality is unimportant, particularly for pipeline systems in which SHI recognition is performed on ASR-generated transcripts. Rather, the divergence between MER and SHI detection performance in the end-to-end setting evaluated here shows that improvements in recording-level SHI detection can occur without corresponding improvements in transcription accuracy. Consequently, transcription accuracy alone is insufficient to characterize recording-level SHI detection performance.

Differences in SHI recognition across categories were consistently observed in the published studies and in our foundation-model comparison. All nine reviewed studies reported category-level differences in performance or data availability, and two trained separate models or adapters for specific SHI categories or category groups. Similarly, no single foundation model in our standardized comparison performed best across all SHI categories, indicating that aggregate performance alone did not capture the relative strengths of individual models. We therefore examined whether these differences could be exploited through type-aware adaptive routing based on category-specific validation performance. The routed model improved macro-F over the best individual model while maintaining comparable micro-F, suggesting that category-level differences across models can provide useful information for model selection and integration. These findings indicate that improvements in overall model capability do not eliminate category-specific performance differences and that these differences can be exploited when combining models for SHI recognition.

The motivation for recording-level clinical speech de-identification extends beyond transcript processing alone. Transcript-based de-identification protects sensitive information represented in text but does not remove SHI that remains directly audible in the source recording. This distinction is increasingly relevant as clinical audio is collected and increasingly reused for documentation, quality assurance, education, research, and AI development. Detecting SHI directly at the recording level can provide the information needed to subsequently remove or transform sensitive speech, supporting secure sharing and safer secondary use of clinical audio. This distinction also has implications for evaluation. Conventional ASR metrics measure transcription accuracy but do not directly measure whether sensitive speech has been correctly identified and localized within the audio or remains undetected. The category-aware, duration-based evaluation framework used in this benchmark therefore evaluates SHI predictions according to both category and temporal overlap (Figure 3). As illustrated in Figure 3, temporally overlapping predictions of the correct SHI category contribute to true-positive (TP) duration, whereas incorrectly predicted intervals and uncovered reference intervals contribute to false-positive (FP) and false-negative (FN) durations, respectively. Category mismatches are penalized even when the predicted and reference intervals overlap in time. By assigning proportional credit according to correctly classified and localized duration, the framework accounts for partial temporal overlap while penalizing missed, overpredicted, and misclassified SHI spans. It therefore captures errors not reflected by transcription accuracy alone and directly quantifies the duration of sensitive speech that is correctly localized, overpredicted, or remains undetected.

**Figure 3.**
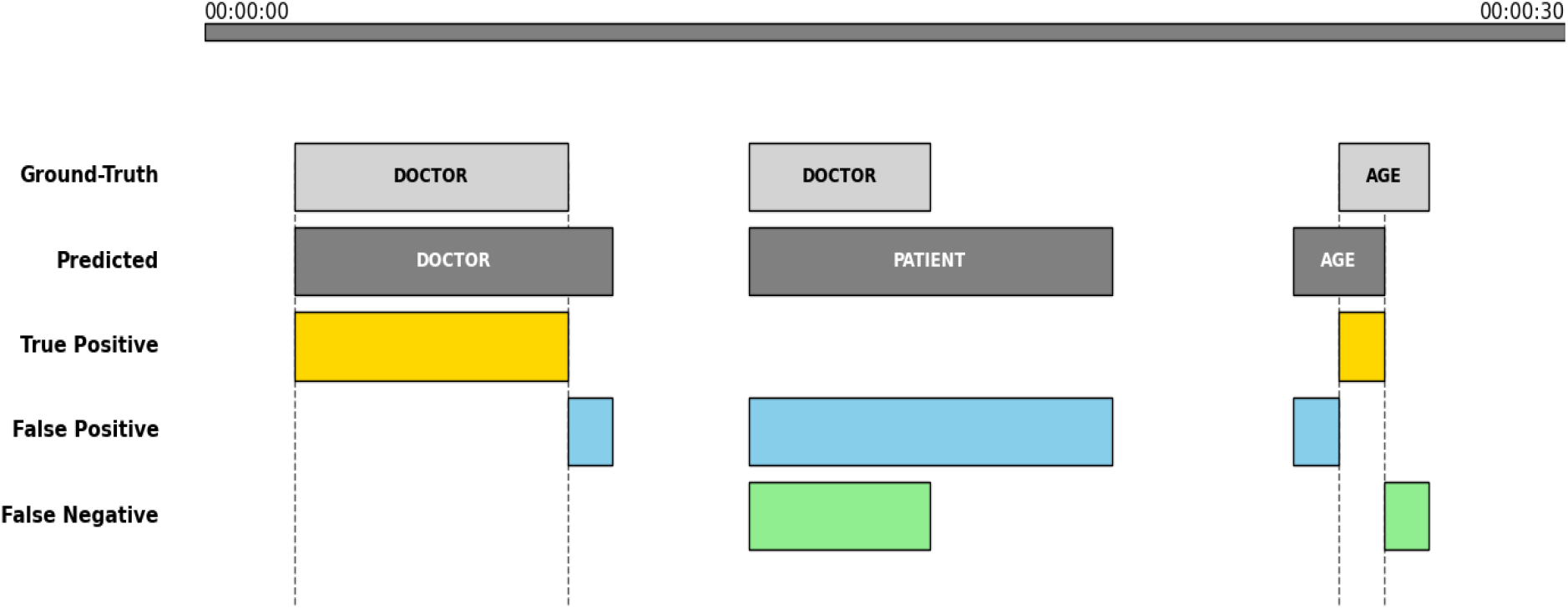
Category-aware duration-based evaluation of temporally localized SHI spans. Predicted SHI intervals were compared with reference annotations along the audio timeline. When a predicted interval and a reference interval had the same SHI category, their overlapping duration was counted as TP; predicted portions extending beyond the reference interval were counted as FP, and uncovered reference portions were counted as FN. When temporally overlapping intervals had different SHI categories, the predicted interval contributed to FP duration for the predicted category, whereas the reference interval contributed to FN duration for the reference category. The framework assigns partial credit based on correctly classified and localized duration while separately penalizing temporal boundary errors and SHI category misclassification.

Recording-level SHI detection is an important step toward clinical audio de-identification, but detecting sensitive information and its temporal boundaries does not by itself remove that information from the source recording. Once sensitive intervals have been identified, they can subsequently be removed, masked, replaced, or otherwise transformed while preserving clinically relevant information in the remaining audio. Evaluation of these transformations will require measures beyond SHI detection, including whether sensitive information remains recoverable after transformation and whether clinically relevant information is preserved. The present benchmark provides a standardized setting for evaluating recording-level SHI detection, whereas evaluation of complete audio de-identification will additionally require assessment of both residual sensitive information and preservation of clinically relevant content.

This study has several limitations. First, the methods identified from the 22 community-developed systems and nine published studies reflect systems developed within the SREDH-AICUP benchmark ecosystem and should not be interpreted as representing all possible approaches to clinical speech de-identification. Technical reports also differed in the level of methodological detail provided, limiting the extent to which individual implementations could be reconstructed or compared. Second, the Mandarin subset was substantially smaller than the English subset of the benchmark. Although synthetic Mandarin speech augmentation improved SHI detection in the end-to-end model, these results were obtained using a specific augmentation procedure and training setting and should not be generalized to other languages, datasets, or augmentation methods without further evaluation. Third, the adaptive routing framework selected and weighted candidate models using category-specific validation performance. Its effectiveness, therefore, depends on the extent to which these category-level performance differences used for routing remain stable in unseen data and should be further evaluated using independent datasets. Fourth, the present experiments were retrospective evaluations rather than prospective evaluations within routine clinical workflows. Further studies using clinical audio from different institutions, recording environments, speaker populations, and clinical settings will be needed to assess robustness and generalizability.

Finally, the present benchmark evaluates recording-level SHI detection but does not evaluate subsequent removal or transformation of detected sensitive speech. Consequently, the present results do not establish whether sensitive information remains recoverable after transformation or whether clinically relevant information is preserved. Future work should evaluate complete audio de-identification systems that remove, mask, replace, or otherwise transform detected SHI and assess both residual sensitive information and preservation of clinically relevant content. Such evaluation will be necessary before performance on recording-level SHI detection can be translated into claims regarding privacy protection or clinical deployment.

## Methods

### Benchmark design and evaluation protocol

The standardized benchmark was designed to evaluate recording-level clinical speech de-identification through two complementary tasks performed on the same collection of clinical speech recordings. Task 1 evaluated automatic medical speech transcription by requiring systems to generate a transcript for each audio recording, whereas Task 2 evaluated recording-level SHI detection by requiring systems to jointly predict SHI categories and their corresponding temporal boundaries (start and end timestamps for each detected span in the audio). Together, these tasks enabled standardized assessment of both speech transcription and recording-level SHI detection under a unified evaluation framework.

Because the two tasks addressed distinct prediction objectives, they were evaluated using separate performance metrics. Task 1 performance was assessed using the MER, which provided a unified measure of transcription accuracy across the English-dominant corpus and its supplementary Mandarin Chinese subset. Task 2 was evaluated using a category-aware, duration-based temporal overlap framework specifically designed for recording-level SHI detection. Unlike conventional entity-level evaluation in text de-identification, this framework jointly assessed SHI classification and temporal localization by directly comparing predicted and reference SHI intervals along the audio timeline.

For each SHI category, the temporal intersection between a predicted interval and a reference interval of the same category was accumulated as TP duration. Portions of a predicted interval that did not overlap a reference interval of the same category were accumulated as FP duration, whereas portions of a reference interval that were not covered by a correctly classified prediction were accumulated as FN duration. Figure 3 illustrates representative scoring scenarios and the accumulation of TP, FP, and FN durations under this framework.

Rather than treating each detected entity as a discrete event, the proposed framework quantified prediction performance using accumulated TP*_c_*, FP*_c,_* and FN*_c_* durations for each SHI category *c*. PRF-scores were therefore calculated from temporal durations rather than entity counts. Macro-F was calculated as the unweighted mean of the category-specific F-scores, whereas micro-averaged measures were calculated after aggregating TP, FP, and FN durations across categories. This category-aware and duration-based formulation enabled the simultaneous evaluation of SHI classification accuracy and temporal boundary localization while directly quantifying correctly localized, overpredicted, and missed SHI intervals across the entire recording.

### Standardized benchmark baseline implementation

To establish standardized reference systems for benchmark evaluation, baseline implementations were developed for both tasks. Task 1 evaluated ASR, whereas Task 2 evaluated recording-level SHI detection using pipeline-based and end-to-end approaches. For both tasks, Whisper served as the reference speech-processing backbone.

For Task 1, standardized Whisper implementations were evaluated across model scales, language configurations, and fine-tuning strategies to establish the reference transcription baseline. Whisper’s predefined control tokens were used to specify the language and, when applicable, the input language. Both the original English-only Whisper models (ranging from Tiny to Medium) and the multilingual Whisper models (including the Large model variants) were evaluated using automatic language identification and manually specified language tokens (Table 1). The multilingual Whisper models were subsequently fine-tuned using bilingual joint training and language-specific independent training (Table 2). Additional implementation details, model configurations, and fine-tuning procedures are provided in the Supplementary Note 1.

For Task 2, two complementary reference baseline strategies were implemented. The official benchmark baseline used an end-to-end architecture based on Whisper without a separate downstream foundation model. This design provided a reference implementation in which speech transcription and SHI detection were performed within a single model.

The end-to-end baseline adapted Whisper’s multitask sequence-generation architecture for joint speech transcription and SHI detection. Consistent with previous studies on joint ASR and speech-based named entity recognition ^46^, tagged transcripts were generated directly from audio. Unlike prompt-conditioned open-type NER, our implementation used a dedicated <|NER|> task token and a fixed XML schema corresponding to the 26 benchmark SHI categories, allowing the model to generate structured SHI annotations directly within the transcript.

In addition to the official end-to-end baseline, standardized pipeline baselines were implemented using the same benchmark data and evaluation protocol. The pipeline combined the independently fine-tuned Whisper large-v3 transcription model with downstream SHI detection models. Three representative downstream models were evaluated under the same pipeline configuration: conventional BIO-based token classification using XLM-RoBERTa-base, and instruction-based SHI generation using the instruction-tuned generative language models Qwen2.5-Omni-7B and Gemma-4-E4B-it-qat-q4_0. For the generative models, SHI entities were represented using category-specific XML-style tags, following previous speech-based named entity recognition work^46^, enabling structured SHI annotation generation through autoregressive next-token prediction ^29^.

To produce standardized recording-level outputs for benchmark evaluation, transcript normalization and forced alignment were applied to the outputs of both the pipeline and end-to-end systems. Model-specific control tokens and XML-style entity tags were removed as applicable while preserving the transcript text for alignment. The normalized transcripts were then aligned to the original audio using the WhisperX forced-alignment framework ^38^ with language-specific alignment models. Word-level timestamps produced by forced alignment were used to reconstruct temporal SHI intervals. For each detected SHI entity, the interval was extended from the start time of the first aligned word to the end time of the last aligned word, producing benchmark outputs consisting of the SHI category, recognized text span, and corresponding start and end timestamps. This procedure ensured that both pipeline-based and end-to-end reference systems produced recording-level outputs in the same format for evaluation under the proposed category-aware, duration-based evaluation framework described above. Additional implementation details, transcript normalization procedure, alignment settings, and model configurations are provided in the Supplementary Note 2.

### Community methodological analysis

To characterize the methods used by competition participants, information was collected from three sources: post-competition participant surveys, official benchmark submissions, and standardized technical reports submitted by participating teams. The participant survey was conducted voluntarily after the competition using an online questionnaire and collected information on model selection, system architecture, and development strategies used during benchmark participation.

Systems were selected for detailed review according to predefined performance criteria rather than competition ranking alone. Teams whose best-performing submissions exceeded the official end-to-end benchmark baseline in either Task 1 or Task 2 were included. Applying these criteria yielded 22 systems with standardized technical reports available for manual review. Participating teams were required to follow a standardized technical report template describing system architecture, training strategy, inference workflow, and implementation details.

Each technical report was manually reviewed to identify the ASR model and language-routing strategy, downstream SHI recognition method, use of data augmentation, category-specific models or adapters, retrieval-based methods, rule-or regex-based processing, temporal alignment method, and fuzzy or approximate span matching. Methods with comparable implementations across systems were grouped, and the number of systems using each method was counted. Method categories were not mutually exclusive because individual systems could incorporate multiple techniques. When a report did not provide sufficient information to determine the specific formulation used for downstream SHI recognition, the system was retained in the review but was not assigned to a more specific SHI recognition category.

Community-wide performance summaries were calculated from the best valid submission from each team. For Figure 2, the displayed community systems comprised the union of the 10 highest-performing teams in Task 1 and the 10 highest-performing teams in Task 2. The standardized benchmark baselines were plotted in the same Task 1-Task 2 performance space for comparison. Because the official competition ranking was determined by the combined Task 1 and Task 2 ranks, Figure 2 was used to visualize the distribution of performance across the two tasks rather than reproduce the official leaderboard.

### Additional benchmark experiments

Based on the review of the 22 community-developed systems and nine published studies, two questions not addressed in the reviewed studies were examined through additional benchmark analyses: (1) whether synthetic Mandarin speech-text augmentation could improve SHI detection in a direct end-to-end model, and (2) we compared contemporary foundation models at the SHI-category level and evaluated whether category-specific differences in model performance could be combined through adaptive routing.

To evaluate Mandarin speech augmentation in the end-to-end setting, we developed a synthetic speech-text augmentation procedure. Training sentences were randomly sampled from the original training corpus in groups of five and provided to gpt-oss-120b to generate additional Mandarin clinical dialogue utterances while preserving XML-based SHI annotations (Supplement Table 2). Speaker diarization and voice activity detection were used to extract reference speech segments prior to synthesis. The generated transcripts were subsequently converted into synthetic speech using BreezeVoice ^47^. During synthesis, XML-based SHI tags were removed from the rewritten transcripts to produce natural target text. Reference speech segments randomly sampled from the original training recordings were used for speaker conditioning during synthesis.

This procedure generated 499 additional Mandarin speech recordings, corresponding to approximately 1.45 hours of synthesized speech and containing 1,948 annotated SHI entities. The synthesized recordings were added to the original training set and evaluated using both bilingual joint training and language-specific independent training with the same end-to-end Whisper architecture and benchmark evaluation protocol described above.

Six contemporary open foundation models were selected for comparison, comprising four general-purpose models, including Qwen2.5-7B, Gemma-2-9B, Mistral-7B, and LLaMA-3.1-8B, and two biomedical models, including OpenBioLLM-8B and Med42-8B. All models were evaluated using the same prompting strategy and standardized XML-based SHI annotation format to minimize differences attributable to task formulation and prompting. For historical comparison, the Pythia-2.8B configurations from our previous transcript-level SHI de-identification study ^29^ were evaluated under the same benchmark protocol.

To investigate whether category-specific differences across foundation models could be combined, we implemented a type-aware adaptive routing framework. Routing was performed independently for each SHI category according to category-specific validation performance. For each SHI category, a per-type performance matrix was first constructed using entity-specific F1 scores on the validation set. For each category, models achieving at least a predefined proportion of the highest-performing model for that entity type were retained as candidate models, from which the top-k models were selected. The selected models were assigned temperature-scaled softmax weights according to their category-specific validation F-scores. To account for differences in the number of training examples across SHI categories, the temperature values were adjusted based on three predefined support levels (common, medium, and rare), resulting in sharper weighting for categories with greater training support and smoother weighting for those with less training support.

Predictions from the selected experts were then combined using weighted voting. For each candidate entity, confidence scores were accumulated across the selected experts according to their routing weights and compared with category-specific acceptance thresholds to determine the final prediction. This procedure allowed multiple models to contribute to each SHI category while assigning greater weight to models with higher category-specific validation performance. All routing parameters were determined using the validation set, and final performance was evaluated under the same category-aware, duration-based benchmark protocol as the individual foundation models.

## Ethics approval

The Human Research Ethics Committee of The University of New South Wales (UNSW Sydney) gave ethical approval for this work (Approval No. HC17749). The datasets had been de-identified prior to access and use by the authors. The corpus used in the SREDH/AICUP 2025 competition is publicly accessible and is subject to approval by signing a data-usage agreement. Access can be requested through the SREDH consortium git profile here (https://github.com/SREDH-Consortium).

## Code availability

More details about the competition and the baseline code can be found here: https://www.codabench.org/competitions/4890/?secret_key=38d92718-cc4d-4907-9c65-c73419671268.

## Data Availability

The corpus used in the SREDH/AICUP 2025 competition is publicly accessible and is subject to approval by signing a data-usage agreement. Access can be requested through the SREDH consortium git profile here (https://github.com/SREDH-Consortium).

http://github.com/SREDH-Consortium/OpenDeID-Corpus

## Acknowledgments

This study was funded by the SREDH/AICUP 2025, which occurred under the auspices of the Competition Guidance Unit within the Department of Information, Technology, and Education at the Ministry of Education (MoE), Taiwan. The Competition Planning Unit of the Artificial Intelligence and Annotation Data Collection Program Office, MoE, Taiwan, provided strategic coordination and oversight. This competition is a collaborative effort involving the Intelligent Systems Laboratory (ISLab) and the Wireless Networking and Mobile Computing Research Lab within the Department of Electrical Engineering, as well as the Department of Applied English at the National Kaohsiung University of Science and Technology; the Machine Learning and Bioinformatics Lab within the Department of Computer Science at the University of Taipei; the Department of Bioinformatics and Medical Engineering at Asia University; and the SREDH Consortium. JJ was funded by the Australian National Health and Medical Research Council (Grant GNT1192469). We acknowledge funding support from the Research Technology Services at the University of New South Wales, Sydney, from ASUSTeK Computer Inc., and from the National Science and Technology Council under grant number NSTC 114-2637-8-992-007. We also thank the Secure Research Environment for Digital Health (SREDH) Consortium Translational Cancer Bioinformatics Working Group for providing access to the OpenDeID corpus.

