## Supplementary for "Benchmarking Recording-Level Clinical Speech De-identification for Privacy-Preserving Clinical Audio"

### Supplementary Information

#### Supplementary Tables

**Supplementary Table 1. Entity-specific F1-scores across representative foundation models for transcript-level SHI recognition.** Entity types with zero F1-scores across all evaluated models are omitted for clarity. The final two rows report overall micro-F1 and macro-F1 scores. Higher values indicate better entity recognition performance. Bold values indicate the highest F1-score for each SHI category among the six contemporary foundation models; ties are retained.

| Type | Qwen 2.5-7B | Gemma-2-9B | Mistral -7B | LLaMa-3.1-8B |
|---|---|---|---|---|
| DOCTOR | 0.8296 | 0.7290 | **0.8557** | 0.8456 |
| DATE | 0.9061 | 0.8961 | 0.9085 | 0.9097 |
| PATIENT | **0.7489** | 0.6928 | 0.7145 | 0.7344 |
| ID_NUMBER | 0.8728 | 0.7637 | 0.8786 | 0.8826 |
| HOSPITAL | 0.8509 | 0.7836 | 0.8328 | 0.8456 |
| MRN. | 0.9080 | 0.8887 | **0.9156** | 0.9114 |
| DEPARTMENT | 0.7410 | 0.6984 | 0.7660 | 0.7617 |
| TIME | 0.8954 | 0.8256 | 0.8892 | **0.8976** |
| CITY | 0.6943 | 0.5894 | **0.7005** | 0.6735 |
| STREET | 0.7725 | 0.7079 | **0.7911** | 0.7715 |
| ZIP | **0.9339** | 0.8145 | 0.8602 | 0.9303 |
| STATE | **0.8450** | 0.6925 | 0.7713 | 0.8348 |
| AGE | 0.8125 | 0.4918 | 0.8718 | 0.8574 |
| FAMILYNAME | 0.0432 | 0.0000 | **0.0999** | 0.0845 |
| SET | 0.5086 | 0.3390 | **0.6188** | 0.5086 |
| DURATION | 0.3466 | 0.1584 | 0.4283 | 0.3564 |
| ORGANIZATION | 0.5756 | 0.5529 | 0.5658 | 0.6586 |
| LOCATION-OTHER | 0.4994 | 0.0000 | 0.5894 | **0.6162** |
| PERSONALNAME | 0.0000 | 0.0297 | 0.1526 | 0.0000 |
| COUNTRY | 0.1888 | 0.0227 | 0.0000 | 0.1888 |
| ROOM | 0.0000 | **0.3442** | 0.0000 | 0.0000 |
| PROFESSION | 0.5579 | **0.8686** | 0.5579 | 0.4670 |
| PHONE | **0.9810** | 0.0000 | 0.0000 | **0.9810** |
| **MICRO F1** | 0.8614 | 0.7979 | 0.8615 | **0.8666** |
| **MACRO F1** | 0.6310 | 0.5169 | 0.5986 | 0.6399 |

| Type | OpenBioLLM-8B | Med42-8B | Pythia-2.8B-ICL | Pythia-2.8B (QLoRA) | Pythia 2.8B Full |
|---|---|---|---|---|---|
| DOCTOR | 0.8408 | 0.8551 | 0.1225 | 0.412 | 0.401 |
| DATE | 0.9098 | **0.9109** | 0.4150 | 0.836 | 0.824 |
| PATIENT | 0.7011 | 0.7426 | 0.1275 | 0.417 | 0.343 |
| ID_NUMBER | **0.8839** | 0.8592 | 0.3200 | 0.608 | 0.586 |
| HOSPITAL | 0.8291 | **0.8758** | 0.0800 | 0.164 | 0.164 |
| MRN | 0.9083 | 0.8757 | 0.4525 | 0.772 | 0.757 |
| DEPARTMENT | 0.7345 | **0.7800** | 0.0575 | 0.249 | 0.272 |
| TIME | 0.8927 | 0.8737 | 0.1050 | 0.502 | 0.475 |
| CITY | 0.6598 | 0.6827 | 0.1200 | 0.323 | 0.321 |
| STREET | 0.7857 | 0.7442 | 0.1100 | 0.246 | 0.217 |

| Type | OpenBioLLM-8B | Med42-8B | Pythia-2.8B-ICL | Pythia-2.8B (QLoRA) | Pythia 2.8B Full |
|---|---|---|---|---|---|
| ZIP | 0.8745 | **0.9142** | 0.2550 | 0.832 | 0.777 |
| STATE | 0.7044 | 0.8170 | 0.2475 | 0.620 | 0.583 |
| AGE | 0.8328 | 0.8610 | 0.2075 | 0.788 | 0.640 |
| FAMILYNAME | 0.0657 | 0.0838 | 0.0000 | 0.000 | 0.000 |
| SET | 0.5086 | 0.5086 | 0.0000 | 0.000 | 0.000 |
| DURATION | 0.4360 | **0.4740** | 0.0000 | 0.118 | 0.111 |
| ORGANIZATION | **0.7356** | 0.7303 | 0.0000 | 0.250 | 0.444 |
| LOCATION-OTHER | 0.3749 | 0.5682 | 0.0000 | 0.000 | 0.000 |
| PERSONALNAME | **0.3627** | 0.1669 | 0.0000 | 0.333 | 0.000 |
| COUNTRY | 0.1888 | **0.2226** | 0.0000 | 0.000 | 0.000 |
| ROOM | 0.2097 | 0.0000 | 0.0000 | 0.000 | 0.000 |
| PROFESSION | 0.5579 | 0.5579 | 0.0000 | 0.000 | 0.000 |
| PHONE | **0.9810** | **0.9810** | 0.0000 | 0.000 | 0.000 |
| **MICRO F1** | 0.8572 | 0.8602 | 0.2735 | 0.5263 | 0.5037 |
| **MACRO F1** | 0.6512 | **0.6559** | 0.1299 | 0.3112 | 0.3008 |

**Supplementary Table 2. Prompt used for text rewriting in the speech-level Mandarin augmentation pipeline.** The prompt instructed the language model to first analyze the contextual, structural, and conversational characteristics of representative clinical dialogues before generating structurally diverse yet clinically coherent Mandarin dialogue utterances while preserving XML-based SHI annotations. Five randomly sampled training utterances were provided as in-context examples for each prompt, and the resulting synthetic dialogues were subsequently converted into speech using the augmentation pipeline described in the Methods.

**Role**
You are an AI expert specializing in clinical dialogue analysis and synthetic medical data generation.

**Task 1: Structural Analysis**
I will provide five medical dialogue utterances annotated with XML tags (e.g., <NAME>, <DATE>, <DOCTOR>).
Analyze these examples and identify the following characteristics:
1. Context
Describe the clinical scenario in which these conversations take place (e.g., emergency department handoff, outpatient consultation, inpatient ward discussion).
2. Logic Flow
Identify the typical conversational structure or sequence of information (e.g., identity verification → symptom description → clinical recommendation).
3. Tone
Analyze the communication style and emotional tone of the speakers (e.g., calm, reassuring, urgent, professional, empathetic).
4. Annotation Patterns
Identify where XML-tagged entities typically appear within the sentences and describe any recurring annotation patterns.

Task 2: Creative Generation

Based on the characteristics identified above, generate 5 entirely new medical dialogue utterances that follow the same overall communication style while remaining semantically and structurally original.
The generated dialogues must satisfy the following requirements:

Originality
- Do not copy or closely imitate the sentence structures from the examples.
- Do not simply replace the entity values while preserving the same syntax.
- Generate genuinely new dialogue patterns.

Scenario Expansion
- Preserve the overall conversational style but create dialogues in different medical specialties (e.g., psychiatry, pediatrics, obstetrics, surgery, oncology, rehabilitation, etc.).

XML Annotation
- Preserve the XML annotation format.
- Insert appropriate XML tags into the generated sentences.
- Ensure that every tagged value matches its corresponding entity type (e.g., <DATE> should contain a valid date expression).

Conversational Naturalness
- Write natural spoken dialogue that resembles authentic clinician–patient conversations.
- Avoid script-like narration such as:
  - “Dr. Smith said…”
  - “The physician instructed…”
- Instead, write only what the speaker would naturally say aloud.

Clinical Realism
- Ensure that the generated conversations are clinically plausible and contextually coherent.

Output Format
- Output only the 10 XML-annotated dialogue sentences.
- Do not include explanations or additional commentary.

Example Input
1. {Sentence 1}
2. {Sentence 2}
3. {Sentence 3}
4. {Sentence 4}
5. {Sentence 5}

**Supplementary Table 3.** Fine-tuning hyperparameters for Whisper transcription model variants.

| Model Size | LoRA Target Modules | Batch | Accumulation Steps | Warmup Ratio | Learning Rate |
|---|---|---|---|---|---|
| large-v3-turbo | q_proj, v_proj | 2 | 8 | 0.1 | 1e-05 |
| large-v3 | q_proj, v_proj, k_proj,out_proj, fc1, fc2 | 2 | 4 | 0.1 | 1e-04 |
| large-v2 | q_proj, v_proj | 2 | 2 | 0.1 | 1e-04 |
| large | q_proj, v_proj, k_proj, fc1, fc2 | 2 | 4 | 0.1 | 1e-05 |
| medium | q_proj, v_proj, k_proj, out_proj, fc1, fc2 | 2 | 4 | 0.1 | 1e-04 |
| small | q_proj, v_proj, k_proj, out_proj, fc1, fc2 | 2 | 2 | 0.1 | 1e-03 |
| base | q_proj, v_proj, k_proj, fc1, fc2 | 2 | 2 | 0.1 | 1e-04 |
| Tiny | q_proj, v_proj, k_proj, out_proj,fc1, fc2 | 2 | 2 | 0.1 | 1e-03 |

**Supplementary Table 4.** Fine-tuning configurations of the standardized benchmark baseline models.

| Hyperparameter | Whisper large-v3 (End-to-end) | XLM-RoBERTa-base | Qwen2.5-Omni-7B | Gemma-4-E4B-it-qat-q4_0 |
|---|---|---|---|---|
| Lora rank | 32 | - | 8 | 8 |
| Lora alpha | 128 | - | 32 | 32 |
| Lora dropout | 0.05 | - | 0.1 | 0.1 |
| Lora target modules | q_proj,v_proj,k_proj, out_proj, fc1, fc2 | - | all-linear | all-linear |
| Batch size | 2 | 32 | 4 | 4 |
| Accumulation S | 4 | 1 | 4 | 4 |
| Optimizer | AdamW | AdamW | AdamW | AdamW |
| Learning rate | 2e-4 | 2e-5 | 2e-4 | 2e-4 |
| Epochs | 3 | 10 | 3 | 3 |
| Warmup Schedule | 50 steps | 0.1 ratio | 0.05 ratio | 0.05 ratio |
| Weight decay | 0.01 | - | - | - |
| Early stop | 2 | 3 | 3 | 3 |

#### Supplementary Notes

##### Supplementary Note 1. Detailed implementation of standardized benchmark baselines

The standardized benchmark baselines were implemented using Whisper's native multitask sequence-generation framework. Whisper controls different speech-processing tasks through predefined control tokens that specify language, task type, and decoding behavior. For conventional speech transcription, the original Whisper transcription format was adopted using the language-specific control token together with the transcription task token. For the end-to-end recording-level sensitive health information (SHI) detection baseline, the original multitask framework was extended by introducing a dedicated <|NER|> task token following the multitask adaptation strategy. This additional control token enabled the model to perform recording-level SHI detection within the same sequence-generation framework while preserving Whisper's original transcription capability.

Recording-level SHI annotations were represented using paired XML tags corresponding to the 26 benchmark SHI categories (e.g., <PATIENT> … </PATIENT>, <DATE> … </DATE>). XML tags were inserted immediately before and after each annotated entity within the reference transcript, allowing SHI extraction to be formulated as a structured sequence-generation task rather than conventional token classification. Because neither the XML tags nor the <|NER|> task token existed in the original Whisper tokenizer, all newly introduced tokens were added to the tokenizer vocabulary before fine-tuning. During inference, the model generated transcripts together with XML-based SHI annotations directly from the input speech, producing structured outputs compatible with the subsequent temporal alignment procedure.

For the comparative pipeline baselines, Whisper was used exclusively for automatic speech recognition (ASR), followed by downstream transcript-level SHI detection. Two representative SHI recognition paradigms were implemented. The encoder-based approach employed XLM-RoBERTa-base using conventional BIO sequence labeling with a token-level classification head. The generative approach employed the instruction-tuned foundation models Qwen2.5-Omni-7B and Gemma-4-E4B-it-qat-q4_0. For these models, transcript-level SHI annotations were reformulated as XML-based generation targets, allowing entity boundaries and entity types to be generated directly through autoregressive next-token prediction rather than sequence labeling. Model-specific chat templates were applied prior to tokenization to conform to the instruction format required by each foundation model.

Whisper transcription models were fine-tuned using parameter-efficient LoRA adaptation. Hyperparameter configurations for different Whisper model variants are summarized in Supplementary Table 3. For transcript-level SHI detection, XLM-RoBERTa-base was trained using full-parameter fine-tuning, whereas Qwen2.5-Omni-7B and Gemma-4-E4B-it-qat-q4_0 were fine-tuned using 4-bit quantization together with LoRA adaptation. Model-specific hyperparameter configurations are summarized in Supplementary Table 4.

For the end-to-end recording-level SHI detection baseline, Whisper-large-v3 served as the backbone model. In addition to LoRA adaptation of the attention modules, the embedding layer and language modeling head were included in fine-tuning to accommodate the newly introduced XML tokens and <|NER|> task token. During preprocessing, transcripts were normalized using Whisper's standard text normalization procedure to maintain consistency with the formatting conventions encountered during model pretraining.

For all experiments, the checkpoint achieving the best validation performance was retained for subsequent evaluation. All experiments were conducted using NVIDIA RTX 4090 GPUs equipped with 24 GB of GPU memory.

#### Supplementary Note 2. Transcript normalization, temporal alignment, and evaluation implementation

To maintain consistency with the formatting conventions learned during Whisper pretraining, all transcripts were normalized using Whisper's standard text normalization procedure prior to model training and temporal alignment. This normalization process removes punctuation and non-linguistic symbols while standardizing abbreviations according to the original Whisper preprocessing pipeline. Applying the same normalization procedure reduced distributional differences between the benchmark corpus and Whisper's pretraining data, thereby providing a consistent textual representation across all experiments.

Because Whisper employs buffered transcription during long-form decoding, transcription timestamps may exhibit temporal drift. To obtain accurate recording-level SHI boundaries, all temporal annotations were generated using the WhisperX forced-alignment framework. Language-specific alignment models were adopted for each language. English recordings were aligned using wav2vec2[1] (WAV2VEC2_ASR_BASE_960H), whereas Mandarin recordings were aligned using XLSR-53 [2] (wav2vec2-large-xlsr-53-chinese-zh-cn). Transcript normalization was first performed according to the corresponding language, after which forced alignment generated word-level timestamps between the transcript and the source recording.

To ensure direct comparability between transcript-mediated pipeline systems and end-to-end recording-level SHI detection systems, the same transcript normalization procedure, alignment models, and timestamp reconstruction strategy were applied consistently across all benchmark baselines and controlled validation experiments. Recording-level SHI intervals were subsequently reconstructed from the aligned word timestamps by assigning the start time of the first aligned word and the end time of the last aligned word within each SHI span.

Task 1 transcription performance was evaluated using the mixed error rate (MER), which provides a unified transcription metric for mixed-language speech containing both English and Mandarin Chinese. Following the benchmark protocol, Chinese text was evaluated at the character level whereas English text was evaluated at the word level. MER was calculated as

$$MER = (S+D+I)/N. \quad (1)$$

where S, D, and I denote the numbers of substitutions, deletions, and insertions, respectively, and N represents the total number of reference tokens (Chinese characters and English words).

Recording-level SHI detection was evaluated using the proposed category-aware duration-based evaluation framework. Unlike conventional entity-level evaluation, which treats each detected entity as a discrete event, the proposed framework accumulates correctly localized and incorrectly localized SHI durations along the audio timeline.

For each SHI category, the temporal overlap between a predicted interval and a reference interval was accumulated as true-positive (TP) duration only when both the SHI category and temporal interval matched. Predicted durations extending beyond the corresponding reference interval were accumulated as false-positive (FP) duration, whereas uncovered portions of the reference interval were accumulated as false-negative (FN) duration.

When predicted and reference intervals temporally overlapped but belonged to different SHI categories, the predicted interval contributed entirely to FP duration for the predicted category, whereas the reference interval contributed entirely to FN duration for the reference category. Consequently, both temporal localization accuracy and SHI category consistency jointly determined the final evaluation score.

Representative examples illustrating correct localization, boundary overprediction, category mismatch, and partial temporal overlap are shown in Figure 3 of the main manuscript.

For each SHI category *c*, precision, recall, and F1-score were calculated from the accumulated TP, FP, and FN durations:

$$Precision_c = \frac{TP_c}{TP_c + FP_c} \quad (2)$$

$$Recall_c = \frac{TP_c}{TP_c + FN_c} \quad (3)$$

$$\text{F1-Score}_c = 2 \cdot \frac{(Precision_c \cdot Recall_c)}{(Precision_c + Recall_c)} \quad (4)$$

Macro-averaged F1-score was computed as the unweighted mean of the category-specific F1-scores across all SHI categories. Micro-averaged precision, recall, and F1-score were calculated after aggregating TP, FP, and FN durations across all SHI categories before metric computation.

Because the proposed evaluation directly measures the duration of correctly localized, missed, and incorrectly predicted SHI speech, it provides a substantially more realistic assessment of recording-level clinical speech de-identification than conventional entity-counting approaches.

Task 1 and Task 2 rankings were calculated independently. Final benchmark ranking was determined by summing the two task-specific ranks, consistent with the ranking procedure adopted in the previous SREDH-AICUP benchmark. This ranking strategy jointly evaluates transcription performance and recording-level SHI detection while preserving the independence of the two benchmark tasks.
